# An early-stage cost and implementation feasibility study of the administration of pro/synbiotics to infants 0-5 months in rural Kenya

**DOI:** 10.1101/2024.12.17.24319142

**Authors:** Beth McCallum, Iwaret Otiti, Florence Achieng, Stephen Allen, Eve Worral

**Affiliations:** Department of Clinical Sciences, Liverpool School of Tropical Medicine, Pembroke Place, Liverpool, UK; Malaria Branch, Kenya Medical Research Institute (KEMRI) Centre for Global Health Research, P.O. Box 1578, 40100 Kisumu, Kenya

**Keywords:** Costing, cost-analysis, programme delivery, implementation, infants, growth, undernutrition, stunting, probiotic, synbiotic

## Abstract

**Background:** Undernutrition underlies approximately 45% of global deaths among children less than five years old, making it one of the most concerning global child health issues. The **PRO**biotics and **SYN**biotics in infants in **K**enya (PROSYNK) trial is assessing whether supervised doses of pro/synbiotics daily for the first 10 days and then weekly to age 6 months (total of 32 doses), has a positive impact on gut health and thereby growth and nutrition. This study is an early-stage cost and implementation feasibility study defining unit costs for the PROSYNK trial and estimating programmatic cost and feasibility of delivering the intervention to infants in rural Kenya.

**Methods:** This provider perspective costing study uses a combination of ingredients approach, activity-based costing and microcosting. First, an empirical cost analysis of the PROSYNK trial was conducted by review of trial documentation and time and motion observations. Next, semi-structured interviews with key informants informed a thematic analysis of implementation feasibility and the development of a theoretical programme structure which formed the basis for estimation of total economic programme costs.

**Results:** The economic cost of delivering the full pro/synbiotics course under trial conditions was measured as $757.32 per participant. Experience gained during PROSYNK and discussions with key informants revealed that it was feasible for the Ministry of Health (MoH) to implement programmatic delivery of the pro/synbiotics, particularly through community-based delivery, without a cold chain and with pro/synbiotic administered directly into infant’s mouths. Incremental economic costs to the MoH of delivering the pro/synbiotic programmatically were estimated to be $9.15 per infant per full course under the base case scenario.

**Conclusion:** Pro/synbiotic administration in early life may be feasible and bear similar costs to existing nutrition interventions. This study will provide policy makers and stakeholders with vital cost and feasibility information to inform effective programmatic implementation in Kenya and similar settings.

## Background

Malnutrition is one of the most concerning global child health issues, with approximately 45% of worldwide deaths among children less than five years old attributable to undernutrition [1]. Children with impaired linear growth, characterised by stunting, also experience suboptimal organ development and cognitive and immune system functioning [2]. Stunting has lifelong and intergenerational socioeconomic consequences, with 43% of children under five in low- and middle-income countries (LMICs) at a greater risk of poverty due to stunting [3]. Children who experience stunting typically earn 20% less income than their non stunted counterparts in adult life [4]. Despite global efforts, the slow decline in the prevalence of stunting highlights the need for further research and continued efforts to combat this issue [5].

PROSYNK (**PRO**biotics and **SYN**biotics in infants in **K**enya) is an intervention trial assessing whether administering pro/synbiotics to newborns daily for the first 10 days and then weekly to age five months (total of 32 doses) in Homa Bay county, western Kenya improves gut health and thereby growth and nutrition [6].

Newborns are randomised to one of four study arms, either a probiotic, one of two synbiotics or control (no intervention). Pro/synbiotics are maintained in a central refrigerator in the Homa Bay Hospital Department of Pharmacy with each dose transported in a cold box by research staff on motorbikes to children’s homes and administered under supervision [7]. Pro/synbiotics are administered either directly into the infant’s mouth or after mixing with sterile water in a medicine cup.

Economic evaluations and implementation feasibility research provide invaluable insights that lay the groundwork for more effective allocation of resources, ultimately leading to enhanced overall health outcomes. This paper provides an early-stage cost analysis of the PROSYNK intervention and considers the feasibility and estimated cost of real-world implementation. It provides context-specific evidence for Homa Bay County where the trial was undertaken.

## Methods

The processes and methods employed to administer pro/synbiotics during the first 0-5 months in infants in Homa Bay County, western Kenya, in the context of a randomised clinical trial have been reported previously [7].

### Trial costing

Data were collected from the PROSYNK trial site, as well as remotely (virtually) from the UK. The remote collection was necessitated by COVID-19 travel restrictions in place at the time. Costing was carried out using a combination of activity-based costing (ABC) and the ingredients approach. This involved identifying activities required to deliver the intervention followed by estimating the quantity and unit cost of the resources (or ‘ingredients’) required for each activity [8].

Data collection consisted of trial documentation review and semi-structured interviews with trial key informants. Initial data collection informed the creation of a comprehensive costing spreadsheet outlining trial activities, subactivities (for activities being microcosted only), and resources used for each of these. Activities were categorised as either trial research costs (TRC) or trial intervention delivery costs (TIDC) by key informants, which allowed research-specific costs to be separated. Activities, subactivities, and ingredients were then shared for review with trial management to check for errors or omissions. Data for each resource used in the trial were outlined on the costing spreadsheet, including a broad cost category (cost category 1), specific cost category (cost category 2), cost type (capital/not-capital), unit type, unit quantity, unit cost, percentage of resources allocated to activity, and useful life of resources.

The percentage of each resource allocated to activities was also defined by trial documentation review and trial personnel interviews (e.g., a study vehicle utilised across several activities may be used 80% of the time for the activity “home visits” and 20% of the time for another activity). Given that personnel time spent delivering the intervention was expected to be a key cost driver, time and motion (TAM) observations were carried out for activities ‘home visits’ and ‘clinic visits’ to facilitate more accurate costing.

Resources were assigned financial costs based on trial expenditure records and confirmed, where possible, with secondary publicly available sources (e.g., manufacturers’ prices of items). Resources were valued using appropriate market rates such as current rent and vehicle purchase costs. Resource values were assigned in the currency used, most often in Kenyan Shillings (KES) and British Pounds (£GBP). KES and £GBP costs were calculated for each item which were subsequently converting to 2021 US dollars ($USD) using respective mean exchange rates for 2021 [9] at the time of study analysis.

In line with World Health Organization (WHO) costing guidance, a standard discount rate of 3% was used for economic costs to account for the opportunity cost of capital assets [10] Discount rate and useful life were used to calculate annualisation factors for capital items. These inputs are outlined for each resource on the costing spreadsheet (available on reasonable request). Annualisation factors were then used to obtain annual economic costs of capital goods.

The trial costing spreadsheet was used to calculate total trial cost which is defined as **the total annualised economic cost of delivering the intervention to all trial participants over a period of one year**. This is presented in $USD. Financial costs and results in other currencies (£GBP and KES) are available as supplementary material.

### Feasibility and Theoretical Programme Structure

Semi-structured interviews with trial field workers, trial management, local healthcare workers and relevant experts and informed a thematic analysis of implementation feasibility and the development of a Theoretical Programme Structure (TPS). Interview guides were developed for each of four key informant types. The themes explored included overall feasibility, health system capacity, delivery and logistics, administration safety and ease, acceptability, potential challenges, supply chain, and stakeholders.

Sections of interview transcripts were coded into the above themes. This allowed themes to be analysed across all transcripts and synthesised to explore themes in detail and identifies aspects of programmatic delivery considered to be the most and least feasible.

Using contributions from key informants, the TPS outlined a potential delivery method with a staffing and resource structure that would be required for the Ministry of Health (MoH) to carry out programmatic implementation of pro/synbiotic delivery across Homa Bay County.

### Estimated programme costs

Scaled-up programmatic costs for the TPS were estimated from the provider (MoH) perspective. Costs were estimated under a base case scenario using the delivery method identified as the most realistic and feasible based on key informant discussions. The base case scenario was defined as follows: initial pro/synbiotic supply and education on how to administer these given to mothers by Community Health Workers (CHWs) (or equivalent role) coinciding with their existing routine postnatal home visits for the first 3 days, with resupply of pro/synbiotics monthly coinciding with existing routine child health clinic visits carried out by clinical officers (or nursing staff). Additional scenario analysis (such as utilising a cold chain or extra consumables for pro/synbiotic administration) is discussed further below. An estimated costing sheet was developed to obtain programmatic cost estimates. Input variables (including those associated with the operational elements outlined above) used to obtain programme cost estimates were derived either from empirical data measured in trial settings, secondary data, or based on assumptions. Input variables are outlined in supplementary material (Supplementary Table 1, Additional File 1). Programmatic costs were estimated using the same methods regarding currency, discount rate, resource valuation and annualisation as outlined above for trial costs.

Cost estimates are presented as economic incremental costs (the additional cost to the MoH above the standard of care (SoC)) of implementing programmatic pro/synbiotic delivery to all infants in Homa Bay County per year in $USD. Unit costs were calculated defined as follows:

Cost per pro/synbiotic = estimated total economic incremental cost above the SoC to the MoH of delivering one pro/synbiotic to one infant in Homa Bay County.

Cost per course = estimated total economic incremental cost above the SoC to the MoH of delivering one full course of pro/synbiotics (total of 32 doses) to one infant in Homa Bay County.

Financial costs and results in £GBP are available as supplementary material.

### Uncertainty and scenario analysis

As in any cost analysis uncertainty and alternative scenarios will affect costs; this was explored in scenario and uncertainty analysis. Input variables for the base case scenario underwent one-way sensitivity analysis to assess the extent of their impact on total programme costs. Programmatic costs were also estimated under different scenarios varying in operational elements that impact the cost and/or feasibility of delivery. Operational elements varied were: the use of a cold chain in pro/synbiotic supply and distribution, and the use of consumables (e.g., medicine cup, sterile water) in administration of pro/synbiotics.

## Results

Trial costs were calculated as shown in Table 1. Detailed trial costing results (including financial costs, reporting in other currencies, and further breakdown of cost categories) are available in Supplementary Table 2, Additional File 1.

**Table 1:** Total Trial Costs.

| **Activity**<br>*Category* | **Total cost**<br>**(\$USD)** | **% of**<br>**grand total** | **% of activity**<br>**category** |
| --- | --- | --- | --- |
| **Management & supervision** | **69,212** | **34.10** | **-** |
| *Labour* | 63,823 | 31.45 | 92.21 |
| *Transport* | 2,733 | 1.35 | 3.95 |
| *Spaces and rent* | 2,071 | 1.02 | 2.99 |
| *Materials and consumables* | 219 | 0.11 | 0.32 |
| *Utilities & bills* | 204 | 0.10 | 0.29 |
| *Tools and equipment* | 163 | 0.08 | 0.24 |
| **Home visit** | **64,514** | **31.79** | **-** |
| *Labour* | 40,085 | 19.75 | 62.13 |
| *Transport* | 13,465 | 6.63 | 20.87 |
| *Materials and consumables* | 9,615 | 4.74 | 14.90 |
| *Spaces and rent* | 828 | 0.41 | 1.28 |
| *Tools and equipment* | 521 | 0.26 | 0.81 |
| **Supply chain** | **37,477** | **18.46** | **-** |
| *Labour* | 14,768 | 7.28 | 39.41 |
| *Transport* | 10,026 | 4.94 | 26.75 |
| *Spaces and rent* | 8,977 | 4.42 | 23.95 |
| *Utilities & bills* | 2,762 | 1.36 | 7.37 |
| *Tools and equipment* | 865 | 0.43 | 2.31 |
| *Materials and consumables* | 80 | 0.04 | 0.21 |
| **Clinic visit** | **13,446** | **6.62** | **-** |
| *Labour* | 9,342 | 4.60 | 69.48 |
| *Materials and consumables* | 1,672 | 0.82 | 12.43 |
| *Spaces and rent* | 1,292 | 0.64 | 9.61 |
| *Tools and equipment* | 1,140 | 0.56 | 8.48 |
| **Viability assessment culture** | **11,125** | **5.48** | **-** |
| *Materials and consumables* | 5,482 | 2.70 | 49.27 |
| *Labour* | 5,439 | 2.68 | 48.89 |
| *Utilities & bills* | 204 | 0.10 | 1.83 |
| **Initiation & training** | **7,190** | **3.54** | **-** |
| *Training* | 3,915 | 1.93 | 54.46 |
| *Initiation* | 2,982 | 1.47 | 41.47 |
| *Utilities & bills* | 204 | 0.10 | 2.84 |
| *Materials and consumables* | 80 | 0.04 | 1.11 |
| *Tools and equipment* | 9 | 0.00 | 0.13 |
| **Grand Total** | **202,964** | **100.00** |  |

### Programmatic Feasibility

Key informants considered that it would be feasible for the MoH to carry out programmatic delivery of pro/synbiotics, particularly through community-based delivery coinciding with routine postnatal care. Detailed thematic analysis results are available (Supplementary Table 4, Additional File 1). The Theoretical Programme Structure was developed as shown in Figure 1.

**Fig. 1:**
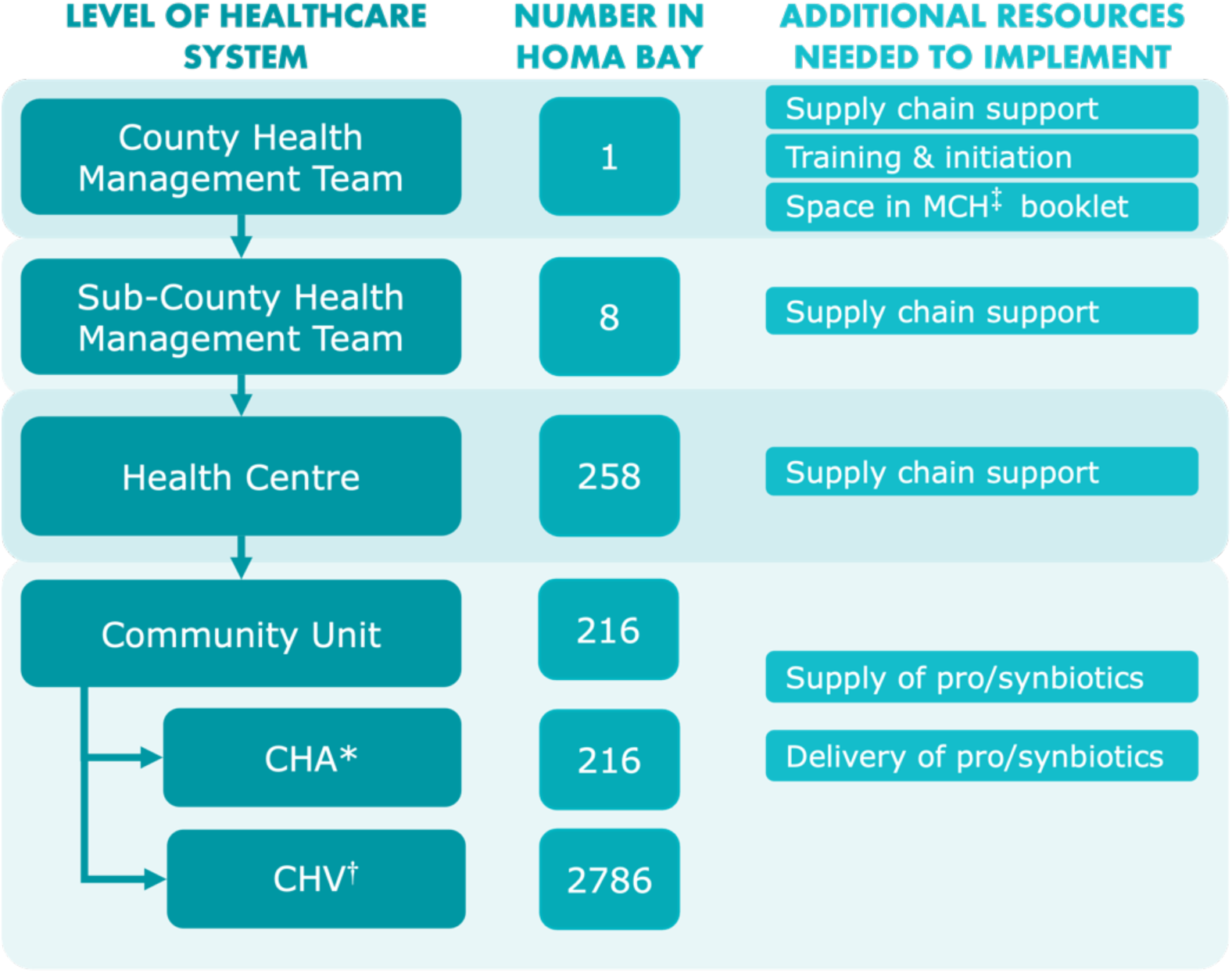
Theoretical Programme Structure. *Community Health Assistant ^†^Community Health Volunteer ^‡^Maternal and Child Health

### Programme cost estimates

Programme cost estimates under base case scenario input variables are outlined in Table 2.

**Table 2:** Estimated programme cost of county-wide delivery.

| Activity<br>*Category* | Total cost<br>(\$USD) | % of grand<br>total | % of activity<br>category |
| --- | --- | --- | --- |
| **Home visit** | **176,870.42** | **46.38** | - |
| *Pro/synbiotic* | 162,758.30 | 42.68 | 92.02 |
| *Labour* | 14,112.11 | 3.70 | 7.98 |
| **Clinic visit** | **170,853.04** | **44.81** | - |
| *Pro/synbiotic* | 162,758.30 | 42.68 | 95.26 |
| *Labour* | 7,910.69 | 2.07 | 4.63 |
| *Materials and consumables* | 184.06 | 0.05 | 0.11 |
| **Supply Chain** | **32,593.28** | **8.55** | - |
| *Supply & distribution* | 32,551.66 | 8.54 | 99.87 |
| *Spaces and rent* | 41.62 | 0.01 | 0.13 |
| **Initiation and training** | **1,003.84** | **0.26** | - |
| *Training* | 569.90 | 0.15 | 56.77 |
| *Initiation* | 433.94 | 0.11 | 43.23 |
| **Grand Total** | **381,320.58** | **100.00** | - |
| **Unit costs** |  |  |  |
| *Cost per course per infant* | **9.15** |  |  |
| *Cost per pre/synbiotic per infant* | **0.29** |  |  |

### Uncertainty and scenario analysis

Sensitivity analysis presents the three most impactful variables considered (Figure 2) and highlights that the cost per pro/synbiotic had, by far, the most significant impact on programme costs. On analysis and costing of varying scenarios, administration method was identified as another very significant factor impacting estimated programme costs. This was due to the differences in consumables required to administer the pro/synbiotic. Lower programme costs were associated with administration directly into the infant’s mouth (as utilised in the base case), as this does not require any additional consumables. This is in contrast to mixing the pro/synbiotic in a container with sterile water before administering, for which costs were estimated using a medicine cup and a small amount of sterile water for each of the 32 doses. The scenario using consumables for the administration of the pro/synbiotic therefore showed an increase in estimated total programme costs, with cost per course increasing from $9.25 to $47.32 (base case using direct administration). Conversely, the scenario utilising a cold chain for supply and distribution of pro/synbiotics resulted only in a modest cost increase, increased the cost per course from $9.25 (base case using ambient supply chain) to $9.61 (with cold chain). Estimated programme costs for these varying scenarios and more detailed base case costing (including financial costs and presenting in £GBP) are available as supplementary material (Supplementary Tables 5-7, Additional File 1).

**Fig. 2:**
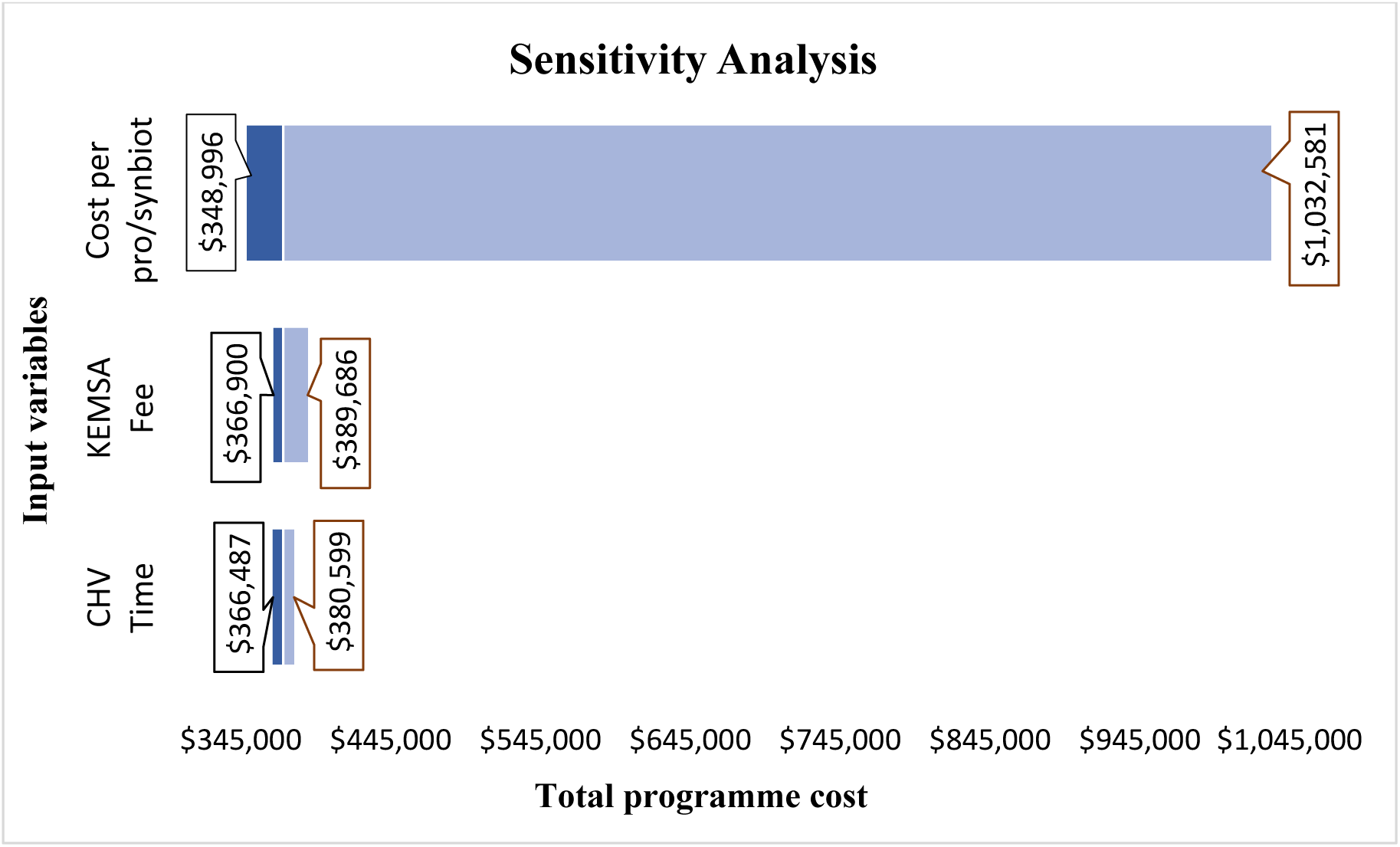
Sensitivity analysis.

## Discussion

We conducted a cost analysis of the PROSYNK trial which showed total trial cost of $202,964.

Interviews indicated that key stakeholders consider the intervention to be feasible and were used to define a TPS with defined staffing. This, along with trial cost data was used to estimate cost of a base case which was estimated to cost $9.15 per course per infant and $0.29 per pro/synbiotic per infant. Scenario analysis revealed that costs increased by 417% with administration via a cup. Use of cold chain increased costs by 5%.

Administration of pro/synbiotics may have significant health benefits in early life [6]. This analysis shows that administration in the community as part of routine child health services in Kenya, without the use of a cold chain and with administration directly into infants mouths, may be affordable with an estimated cost per 6-month course (32 dose) per infant of USD $9.15.

The results of this study will guide future research (ideally with embedded cost-effective analysis) and offer policymakers and stakeholders valuable insights into cost and feasibility for potential implementation, informing efficient intervention design. Decentralisation in Kenya also indicates that the results of this study will be especially useful to Homa Bay County, as counties have their own budgets and can allocate them to relevant priorities.

Homa Bay County has a dispersed rural population and poverty rates above the national average [11, 12]. Therefore, it can be logistically and financially difficult for individuals to attend facilities [13]. For this reason, key informants considered that community-based delivery led by CHVs (without the use of a cold chain) was the most feasible. The use of CHVs for similar interventions has been shown to be cost-effective and improve healthcare coverage, particularly in rural areas [14–19] such as Homa Bay County. The role of CHVs in pro/synbiotic delivery would be expected to include delivery of an initial supply of pro/synbiotics and teaching on how to administer these coinciding with existing routine postnatal home visits for the first 3 days (as was utilised in the estimation of programme costs).

The point estimates for economic and financial programme costs were $381,320.58 and $359,171.84, respectively. Pre-existing resources already owned by the MoH used in programme delivery, such as clinic space and staff, have no financial cost and bare only economic costs; these resources account for much of the difference between economic and financial costs. Economic costs include the opportunity cost of utilising pre-existing resources for programmatic delivery and, therefore, are more representative of the resources (rather than finances) needed, as it does not assume pre-existing resources to be “free”.

Sensitivity analysis showed that total programme costs were very sensitive to the cost per pro/synbiotic. Cost per pro/synbiotic is a particularly important variable to consider, particularly as the ongoing PROSYKN trial is currently investigating three distinct formulations (each bearing different costs). This highlights the importance of further investigation, if the intervention is shown to be effective, to determine which pro/synbiotic formulation would be used. This also highlights how reductions in cost when buying in bulk for programmatic delivery has the potential to significantly reduce programmatic costs. Other variables, such as healthcare worker time spent on pro/synbiotic delivery, fee for warehousing and distribution, and costs of training and initiation, were also examined. However, these factors had a very modest impact on total estimated costs compared to the cost per pro/synbiotic.

A substantial difference in estimated program costs were observed between different administration methods. Specifically, the scenario utilising consumables (e.g., mixing cups and sterile water which was included in the PROSYNK trial) to administer the pro/synbiotic resulted in a large increase in costs from the base case scenario, where the pro/synbiotic would be administered directly into the infant’s mouth. Direct administration not only bares lower estimated costs but was also considered to be highly feasible and realistic. Experience from the PROSYNK trial indicated that direct administration to infants was preferred by many mothers, suggesting its potential adoption as the primary delivery method outside of a trial context.

Although less impactful on estimated costs than some of the other variables considered, the use of a cold chain was highlighted during key informant interviews as an operational element which could significantly reduce the feasibility of programmatic implementation. Additional costs incurred when utilising a cold chain were due to capital purchases (fridges, cool boxes) and an increased fee from Kenya Medical Supplies Agency (KEMSA, the government medical logistics provider contracted by county governments to delivery medical commodities) for procurement, warehousing, and distribution under a cold chain. However, stability testing of the pro/synbiotics at ambient temperatures in Kenya indicate that a cold chain would not be required (K. Otieno; personal communication).

Unit costs of existing interventions may provide context for nutrition spending in Kenya. Some financial unit costs (per beneficiary per year) in Kenya are reported by The World Bank [20] as follows: vitamin A supplementation ($0.44), therapeutic zinc supplementation with ORS ($1.34), multiple micronutrient powders ($2.00), public provision of complementary food for prevention of moderate acute malnutrition ($47.99), and treatment of severe acute malnutrition ($83.32). The programmatic financial unit cost of delivering one full course of pro/synbiotics to one infant was estimated to be $8.62 under base case conditions in our study (this was $9.15 when considering economic costs, which the world bank examples do not). This is within a similar cost range to these existing interventions.

Cost-effectiveness analysis is the preferred economic analysis method to evaluate interventions but, as the PROSYNK trial is ongoing, the effectiveness of the intervention is not yet known, precluding a cost-effectiveness analysis. Further, cost estimates presented for the PROSYNK intervention are based on costs of undertaking a limited research project under trial conditions. Due to highly regulated research conditions trial costs are not representative of routine intervention delivery costs, and our estimated costs did not consider potential reduced costs when buying in bulk (for both pro/synbiotic and consumables) for programmatic delivery. However, our methodology used key informant interviews to try and adapt these data to a realistic programmatic delivery costing.

This study took the provider perspective. A societal perspective, which considers the financial and economic cost to participants, would be best practice. Many economic analyses in LMICs highlight important opportunity costs incurred by beneficiaries of existing community-based nutrition programmes [13]. Therefore, it would be crucial to also consider costs to beneficiaries in future research and if the intervention were to be delivered programmatically (particularly when considering and comparing possible delivery mechanisms).

To overcome these limitations, we recommend an embedded cost-effective analysis in further studies that take forward the findings from the PROSYNK trial.

## Conclusion

Prioritising effective implementation of interventions to improve nutrition that have been proven to be affordable and feasible using context-specific data grounded in local healthcare systems is critical to ensuring improved health outcomes. Despite methodological limitations that could be addressed in follow-on studies, these results suggest that pro/synbiotic administration in early-life is an affordable intervention when delivered at scale. Opting for community-based delivery without a cold chain and pro/synbiotic administration directly into infants mouths would minimise provider costs and enhance feasibility.

## Data Availability

All data produced in the present study are available upon reasonable request to the authors

## List of Abbreviations

ABC: Activity-based costing
CHA: Community health assistant
CHV: Community health volunteer
GBP: Great British Pounds
KEMRI: Kenya Medical Research Institute
KES: Kenyan Shillings
LMICs: Low- and middle-income countries
LSTM: Liverpool School of Tropical Medicine
MCH: Maternal and Child Health
MoH: Ministry of Health
PROSYNK: PRObiotics and SYNbiotics in infants in Kenya
SoC: Standard of Care
TAM: Time and motion
TIDC: Trial intervention delivery costs
TPS: Theoretical programme structure
TRC: Trial research costs
USD: US Dollars
WHO: World Health Organization
KEMSA: Kenya Medical Supplies Authority

## Declarations

### Ethics approval and consent to participate

The PROSYNK trial, and subsequently this study, ensured compliance with the Declaration of Helsinki: Ethical Principles for Medical Research Involving Human Subjects (World Medical Association, 2013) and all applicable local regulatory requirements in Kenya. Ethical approval was granted by ethics committees at the Liverpool School of Tropical Medicine (LSTM; 20(09)) and Kenya Medical Research Institute (KEMRI; KEMRI/SERU/CGHR/320/3917). There were no deviations from the approved study protocol.

### Consent for publication

N/A

### Availability of data and materials

The datasets supporting the conclusions of this article are available from the corresponding author on reasonable request.

### Authors Contributions

SA and EW conceptualised and all authors designed the study. BM and FA collected data. BM analysed the data with supervision from EW. BW wrote the first draft, EW and SA provided important edits. All authors read and approved the final manuscript.

## Acknowledgements

We thank the key informants for their valuable insights into the operational aspects of pro/synbiotic delivery in communities in Kenya.

## Footnotes

**Supplementary Table 1:** Input variables for estimated programme costs.

| Input Variable | Lower | Baseline | Upper | Justification | Source |
| --- | --- | --- | --- | --- | --- |
| Number of infants in Homa Bay County | 36371 | 40412 | 44453 | Upper value represents the reported population of infants (children aged 0-12 months) in Homa Bay County. Baseline value accounts for total infant mortality, which is considered a more realistic baseline than the total reported population. Lower value accounts for total neonatal (children aged 0-28 days) mortality which is higher than total infant mortality. | Secondary Data [Republic of Kenya MoH, 1, IHME, 2, CICF, 3] |
| Number of nurses in Homa Bay | 194 | 216 | 238 | Baseline value of nurses from secondary data. Upper value accounts for 10% enrolment, lower value accounts for 10% attrition | Secondary data [Republic of Kenya MoH, 1] and assumptions for upper and lower values |
| Number of CHA*s in Homa Bay | 0.31 | 0.31 | 0.31 | Baseline value of CHAs from secondary data. Upper value accounts for 10% enrolment, lower value accounts for 10% attrition | Secondary data [Republic of Kenya MoH, 4] and assumptions for upper and lower values |
| Number of CHV<sup>†</sup>s in Homa Bay | 2507 | 2786 | 3065 | Baseline value of CHVs from secondary data. Upper value accounts for 10% enrolment, lower value accounts for 10% attrition | Secondary data [Republic of Kenya MoH, 4] and assumptions for upper and lower values |
| Number of pro/synbiotics per full course | 22 | 32 | 42 | Baseline value is the total number of supplements in one course used in trial settings. Upper and lower values are assumptions. | Empirical data and assumptions |
| Time spent on IP per HCW<sup>§</sup> per year (hours) | 216 | 240 | 264 | Baseline value is based on the participant-facing time of personnel under trial settings for the first IP dose only. Patient-facing personnel time for subsequent doses were not included as feasible programmatic delivery was identified as unsupervised administration following initial education and IP distribution. Upper and lower values are assumptions of 10% fluctuation in this figure | Assumptions based on empirical data |
| Non-IP consumable cost per supplement (£GBP) | 0.12 | 0.21 | 0.47 | Upper value is the cost of all non-IP consumables required at the point of IP administration in trial settings. Baseline value removes COVID-19 PPE¶ consumables cost as measured in trial settings. Lower value removes the cost of consumables that may or may not be required in real-world settings (e.g., no mixing cup or sterile water). | Empirical data |
| Training cost (£GBP) | 931.34 | 1862.67 | 2822.233624 | All training costs are annualised financial costs. Upper value is total training costs measured in the trial, baseline is 2/3 of this and lower is 1/3. Trial training costs were not considered to be the baseline as training costs were aggregated and therefore will include research-specific aspects | Assumptions based on empirical data |
| Initiation cost (£GBP) | 658.80 | 1356.51 | 2055.325328 | All initiation costs are annualised financial costs. Upper value is trial initiation costs, baseline is 2/3 of this and lower is 1/3. Trial initiation costs were also aggregated and therefore not considered to be the baseline due to likely inclusion of research-specific aspects | Assumptions based on empirical data |

\* Community Health Assistant † Community Health Volunteer ‡ Investigational Product § Healthcare Worker ¶ Personal Protective Equipment

**Supplementary Table 2:**
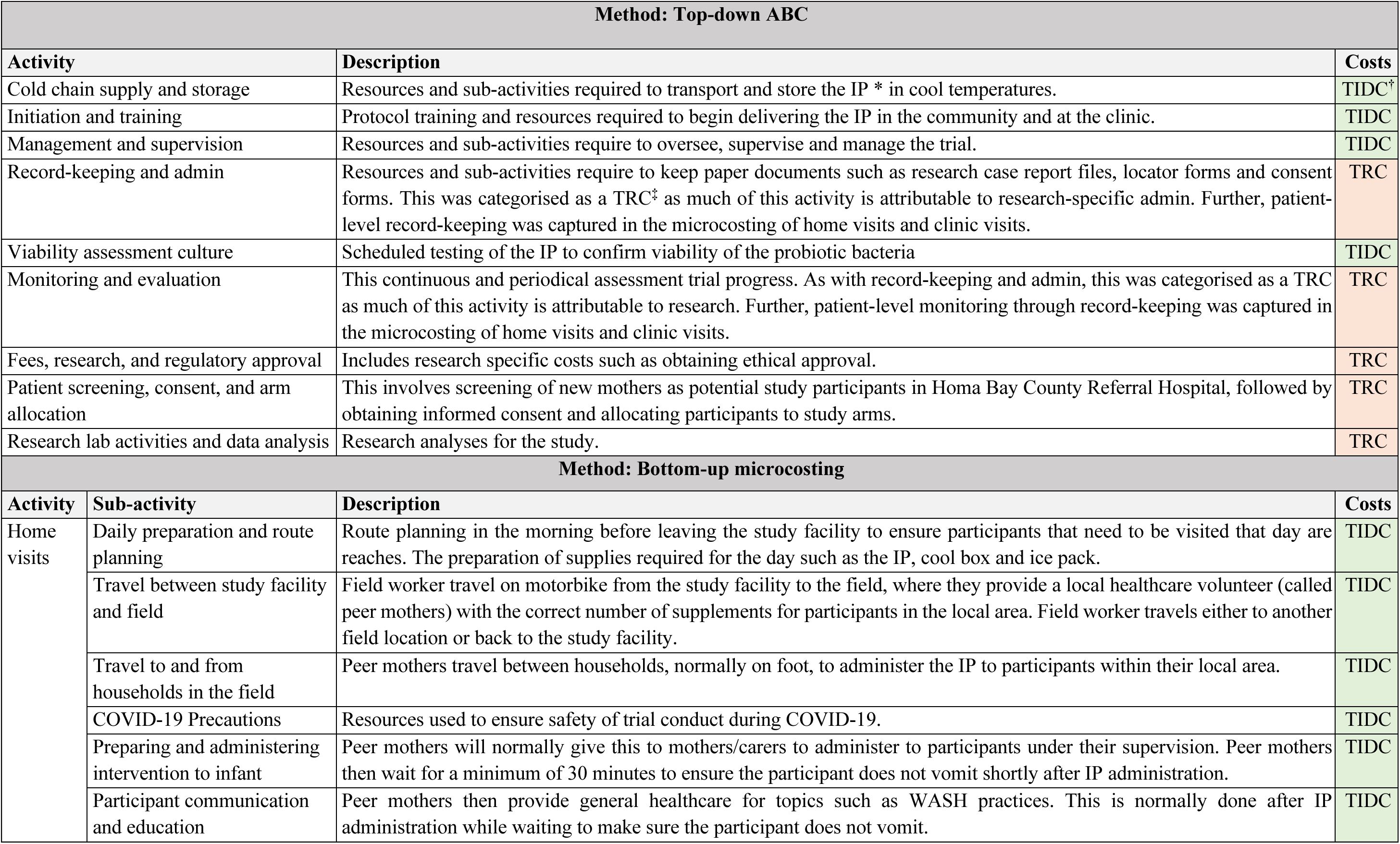
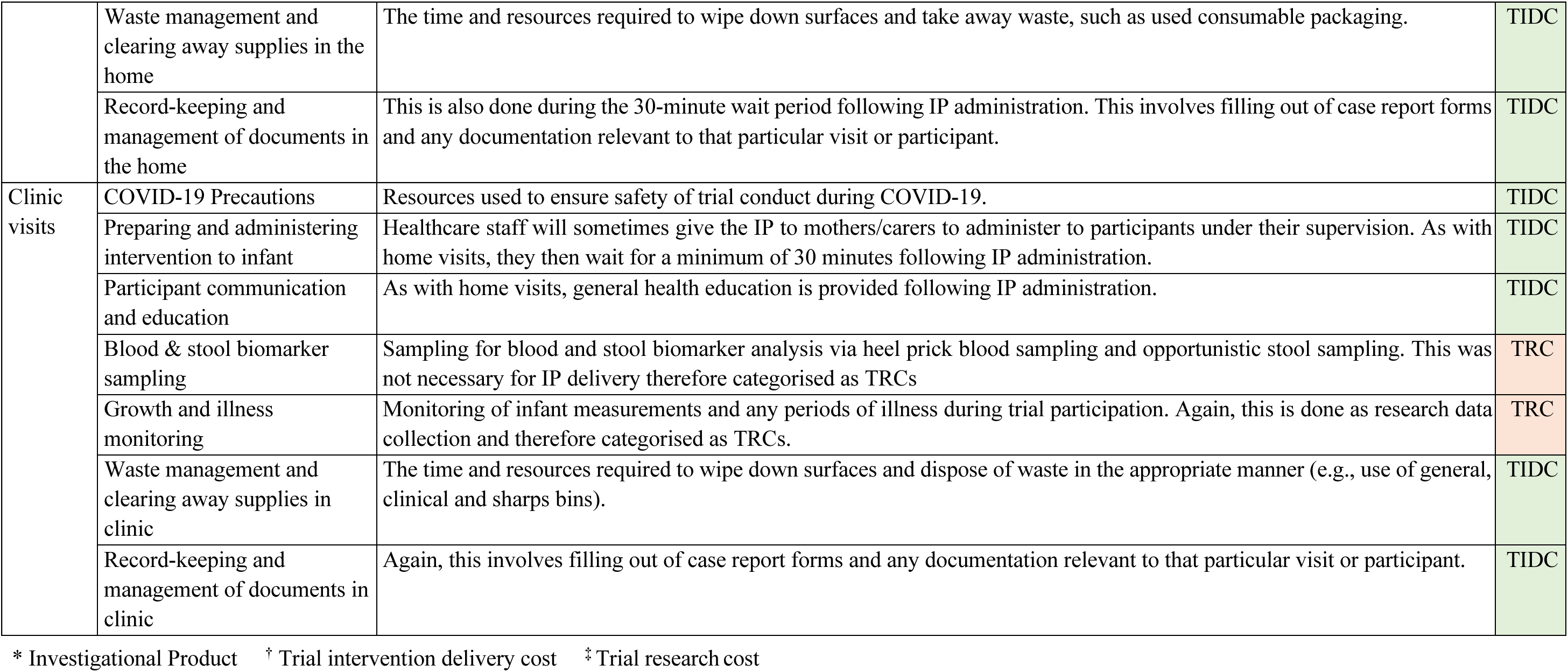
Trial Activities, Costing Methods, and Categorisation.

**Supplementary Table 3:** Trial cost categories, all currencies.

| **Cost category**<br>*Sub-category* | **% total**<br>**economic cost** | **% total**<br>**financial cost** | **Economic**<br>**cost (\$USD)** | **Financial cost**<br>**(\$USD)** | **Economic**<br>**cost (£GBP)** | **Financial cost**<br>**(£GBP)** | **Economic**<br>**cost (KES)** | **Financial cost**<br>**(KES)** |
| --- | --- | --- | --- | --- | --- | --- | --- | --- |
| **Labour** | **65.75** | **72.09** | **133,458** | **133,228** | **96,200** | **96,034** | **14,500,343** | **14,475,343** |
| *Management & supervisory* | 35.07 | 38.52 | 71,183 | 71,183 | 51,311 | 51,311 | 7,734,124 | 7,734,124 |
| *Field workers* | 20.44 | 22.45 | 41,486 | 41,486 | 29,904 | 29,904 | 4,507,517 | 4,507,517 |
| *Medical personnel* | 5.84 | 6.42 | 11,857 | 11,857 | 8,547 | 8,547 | 1,288,294 | 1,288,294 |
| *Support staff* | 4.29 | 4.71 | 8,701 | 8,701 | 6,272 | 6,272 | 945,408 | 945,408 |
| *Pharmacy staff* | 0.11 | 0.00 | 230 | 0 | 166 | 0 | 25,000 | 0 |
| **Transport** | **12.92** | **5.84** | **26,223** | **10,794** | **18,902** | **7,781** | **2,849,143** | **1,172,821** |
| *Fuel, insurance & maintenance* | 5.28 | 5.78 | 10,717 | 10,686 | 7,725 | 7,703 | 1,164,381 | 1,161,064 |
| *Cars* | 4.15 | 0.00 | 8,416 | 0 | 6,067 | 0 | 914,420 | 0 |
| *Motorbikes* | 3.44 | 0.00 | 6,972 | 0 | 5,026 | 0 | 757,507 | 0 |
| *Travel aids* | 0.06 | 0.06 | 118 | 108 | 85 | 78 | 12,835 | 11,757 |
| **Materials and consumables** | **8.45** | **9.28** | **17,146** | **17,146** | **12,360** | **12,360** | **1,862,967** | **1,862,967** |
| *Clinical consumables* | 2.76 | 3.03 | 5,601 | 5,601 | 4,037 | 4,037 | 608,552 | 608,552 |
| *Lab consumables* | 2.57 | 2.82 | 5,210 | 5,210 | 3,755 | 3,755 | 566,045 | 566,045 |
| *Pro/synbiotic* | 1.25 | 1.37 | 2,540 | 2,540 | 1,831 | 1,831 | 275,968 | 275,968 |
| *PPE* | 1.14 | 1.25 | 2,306 | 2,306 | 1,662 | 1,662 | 250,534 | 250,534 |
| *Office supplies* | 0.50 | 0.55 | 1,011 | 1,011 | 729 | 729 | 109,833 | 109,833 |
| *Documentation* | 0.24 | 0.26 | 479 | 479 | 345 | 345 | 52,035 | 52,035 |
| **Spaces and rent** | **6.49** | **6.15** | **13,168** | **11,373** | **9,492** | **8,198** | **1,430,727** | **1,235,699** |
| *Storage space* | 4.42 | 3.88 | 8,974 | 7,179 | 6,468 | 5,175 | 975,000 | 780,000 |
| *Non-clinical room* | 1.43 | 1.57 | 2,902 | 2,902 | 2,092 | 2,092 | 315,327 | 315,299 |
| *Clinical room* | 0.64 | 0.70 | 1,292 | 1,292 | 931 | 931 | 140,400 | 140,400 |
| **Training** | **1.93** | **1.94** | **3,915** | **3,586** | **2,822** | **2,585** | **425,400** | **389,666** |
| **Utilities & bills** | **1.66** | **1.83** | **3,374** | **3,374** | **2,432** | **2,432** | **366,570** | **366,570** |
| **Initiation** | **1.47** | **1.49** | **2,982** | **2,749** | **2,149** | **1,981** | **323,944** | **298,667** |
| **Tools and equipment** | **1.33** | **1.39** | **2,698** | **2,567** | **1,945** | **1,850** | **293,127** | **278,904** |
| **Grand Total** | **100** | **100** | **202,964** | **184,818** | **146,301** | **133,221** | **22,052,221** | **20,080,638** |

**Supplementary Table 4:** Thematic analysis results.

| Thematic Analysis Results | Quotations |
| --- | --- |
| *Theme: Overall Feasibility* |  |
| All respondents felt that overall, it would be feasible for the MoH* to deliver the IP<sup>†</sup> in Homa Bay County. However, several potential considerations and challenges were noted, as discussed in this table. | *“At county level it will be a success because we value our babies so much and we have good nurses here who can work on it and follow up on it and make sure that it is a success.”* |
| *Theme: Health System Capacity* |  |
| All respondents believed that the IP could be delivered with existing local MoH health system capacity in Homa Bay County. | *“The health system has the storage facilities enough to accommodate supplies and the technical staff to manage the supplements to the infants.”* |
| *Theme: Administration safety & ease* |  |
| All respondents believed that the IP was safe to give to mothers and infants unsupervised, however it was noted that this would not be appropriate in research settings as it would not allow for rigorous quality control.<br>Trial personnel stated that they have not witnessed any serious adverse events following IP administration. All but one individual felt that the supplement was easy to administer and that this could be done by mothers, provided they first receive appropriate training. One individual felt that mothers should not administer the supplement on their own as they may store and prepare it incorrectly under unsanitary conditions.<br>The method of IP administration was noted to change with increasing age of infants. During the first days and weeks of life, it was necessary for the IP to be mixed in a medicine cup with a small volume of sterile water or expressed breast milk before being administered. After the first few weeks of life, the IP could be administered in powder form directly into the participants mouth, which was noted to be the easiest and quickest method. | *“We’ve not encountered serious events surrounding supplement administration at home. So, in my thoughts it’s safe.”*<br>*“Mothers can be taught how to give the IP to infants, so I don’t think there are any great concerns.”*<br>*“We’ve not elicited any big challenge regarding the administration of this supplement.”* |
| *Theme: Acceptability* |  |
| All respondents noted that mothers and carers were generally accepting of the IP, with most stating that local communities have been enthusiastic about receiving the supplement. One respondent stated that some community members who are not involved in the trial have shared concerns regarding the IP with the mothers/carers of participants, which has led to them considering withdrawal from the trial. Several respondents noted that community mobilisation would be important for general | *“I have not heard any mother complaining about this food, they really love this food.”*<br>*“So far, it also appears that the community accepts it. Very few numbers of our* |
| acceptability if the MoH were to implement IP delivery in Homa Bay. This will not be discussed further as detailed qualitative analysis of IP acceptability has been carried out as a separate study. | *participants are declining taking the supplements."* |
| *Theme: Potential Challenges* |  |
| The two challenges under trial conditions noted most frequently by trial personnel were conduct of the trial during the COVID-19 pandemic, and frequent MoH staff strikes. Other challenges included frequent laboratory and pharmacy supply shortages and the logistics require to plan home visits for all participants.<br>Potential challenges noted by respondents regarding programmatic implementation of the IP included the following:<br>• Poor sanitation practices in the community<br>• Lack of community acceptability<br>• Poor uptake and coverage of previous evidence-based interventions<br>• Inability to produce supplements in-country<br>• Lack of healthcare regulation and management structures at community level<br>• Lack of accessibility at hospital level | *"The challenges that we have faced, number one, is conduct of this trial in the wake of COVID-19."*<br>*"There are frequent healthcare worker strikes, meaning that recruitment from our delivery unit is a challenge. That has happened a lot."*<br>*"There will be great need for sensitization and awareness because I feel that there is little information, or complete lack of information, about probiotics and synbiotics in Kenya and about what they are doing. So, once the information is spread out and the supporting data about its safety is available to the public, then I think we can go a long way."*<br>*"Already we have many evidence-based interventions for new-born and child health that are shown to be cost effective, but they haven't yet been implemented. Part of it is a poor uptake, but also most importantly poor coverage."* |
| *Theme: Delivery* |  |
| Suggested delivery methods included the following:<br>• If cold chain requirement is confirmed, then the IP could be delivered alongside existing local vaccine cold chain systems. Mothers/carers could be given a supply of supplements when attending routine clinic visits.<br>• Establish an IP supply to level I or II (community) facilities. IP could be collected and distributed (on foot) by CHVs<sup>‡</sup>. Mothers could also collect the IP (on foot) directly from dispensaries if appropriate.<br>Community care (level I-II) was considered the most appropriate and feasible method. Both methods require initial education and IP distribution by MoH staff followed by unsupervised administration by mothers/carers. | *"Mothers would bring the child for growth monitoring and get supplement refills in such settings. That would be the most ideal setting for upward scale."*<br>*"We have dispensaries in our villages which means the IPs will be closer to the community. It would be very feasible. The MoH has been using the CHVs as their gatekeepers, they help them do every intervention they have. This too can be done by CHVs."* |
| *Theme: Accessibility* |  |
| Respondents who commented on accessibility noted that community-based IP delivery would increase this factor. | *"Accessibility is always an issue. You will find that mothers who need to go to the hospital maybe for vaccination or medical concerns but getting there will always be a challenge."* |
| *Theme: Regulation* |  |
| This was noted by several respondents as an important consideration regarding programmatic implementation of the IP. Respondents believed that regulation of supplement administration, supply and quality would be an important factor in ensuring success. One method suggested to potentially improve IP regulation was the addition of one page dedicated to the monitoring of IP administration in pre-existing Kenyan MCH<sup>§</sup> booklets MCH booklets are already used throughout Kenya to monitor the health of mother and child, including the status of other preventative interventions such as vaccinations and vitamin A supplementation. Respondents felt this would allow mothers/carers and healthcare workers to keep track of progress through the entire course of IP administration. | *“Systems to monitor how it's being delivered, who's delivering it, where it's being delivered, and at what time points, and ensuring that mothers have received it and taken it, I think these systems need to be strengthened.”* |
| *Theme: Supply Chain* |  |
| It was noted by several respondents that community-based delivery would not be as feasible if use of a cold chain was proven to be absolutely necessary for transport and storage of the IP, as most households do not have access to cold equipment such as refrigerators. Several respondents stated that viability of the IP when kept at room temperature should be assessed to confirm whether cold chain supply is necessary. In summary, respondents believed that a community level delivery model without the use of a cold chain would be the most feasible option for the MoH to implement programmatic IP delivery. However, utilising local vaccine cold chain supplies was noted as a potential solution. | *“The issue is the cold storage, that would be the major issue. But I think if we go the vaccine way, it will not be a challenge.”* |
| *Theme: Stakeholders* |  |
| Important stakeholders in IP delivery were thought to be the following:<br>● National Kenyan MoH<br>● County health officials of both child health and nutrition sectors<br>● Entire family (carers) of the infant<br>● Appropriate national bodies such as the Kenyan Paediatric Association | *“I think the county health officials are really key because they not only oversee the activities and budgets for the health facilities, but they also oversee what's happening in the community.”* |
| *Theme: Trial Outcome Opinions* |  |
| Many respondents shared their opinions on the outcome of the trial, with all such comments being positive. Respondents believed that the IP would benefit infants and their families. | *“I recommend that the government adopts these supplements as one of the remedies put into place for each and every child born.”*<br>*“The children on the supplements are very healthy, and the mothers are very happy. It will really improve the overall health and development of infants and reduce infant mortality in this country.”* |

\*Ministry of Health
† Investigational Product
‡ Community Health Volunteer
§Maternal and Child Health

**Supplementary Table 5:** Estimated programme cost of county-wide delivery: base case scenario.

|  | Estimated Programme Costs: Base Case Scenario |  |  |  |  |  |
| --- | --- | --- | --- | --- | --- | --- |
| **Cost Drivers**<br>*Activity* | **% total economic cost** | **% total financial cost** | **Economic Cost (\$USD)** | **Financial Cost (\$USD)** | **Economic Cost (£GBP)** | **Financial Cost (£GBP)** |
| **Pro/synbiotic** | **85.37** | **90.63** | **325,517** | **325,517** | **234,640** | **234,640** |
| **Supply & distribution** | **8.55** | **9.06** | **32,593** | **32,552** | **23,494** | **23,464** |
| **Labour** | **5.78** | **0.00** | **22,023** | **0** | **15,875** | **0** |
| *Home visit* | 3.70 | 0.00 | 14,112 | 0 | 10,172 | 0 |
| *Clinic visit* | 2.07 | 0.00 | 7,911 | 0 | 5,702 | 0 |
| **Initiation and training** | **0.26** | **0.26** | **1,004** | **920** | **724** | **663** |
| **Materials and consumables** | **0.05** | **0.05** | **184** | **184** | **133** | **133** |
| **Grand Total** | **100.00** | **100.00** | **381,321** | **359,172** | **274,865** | **258,900** |
|  | **Unit costs** |  |  |  |  |  |
| *Cost per course* | - | - | **9.25** | **8.62** | **6.60** | **6.18** |
| *Cost per pre/synbiotic* | - | - | **0.29** | **0.27** | **0.21** | **0.19** |

**Supplementary Table 6:**
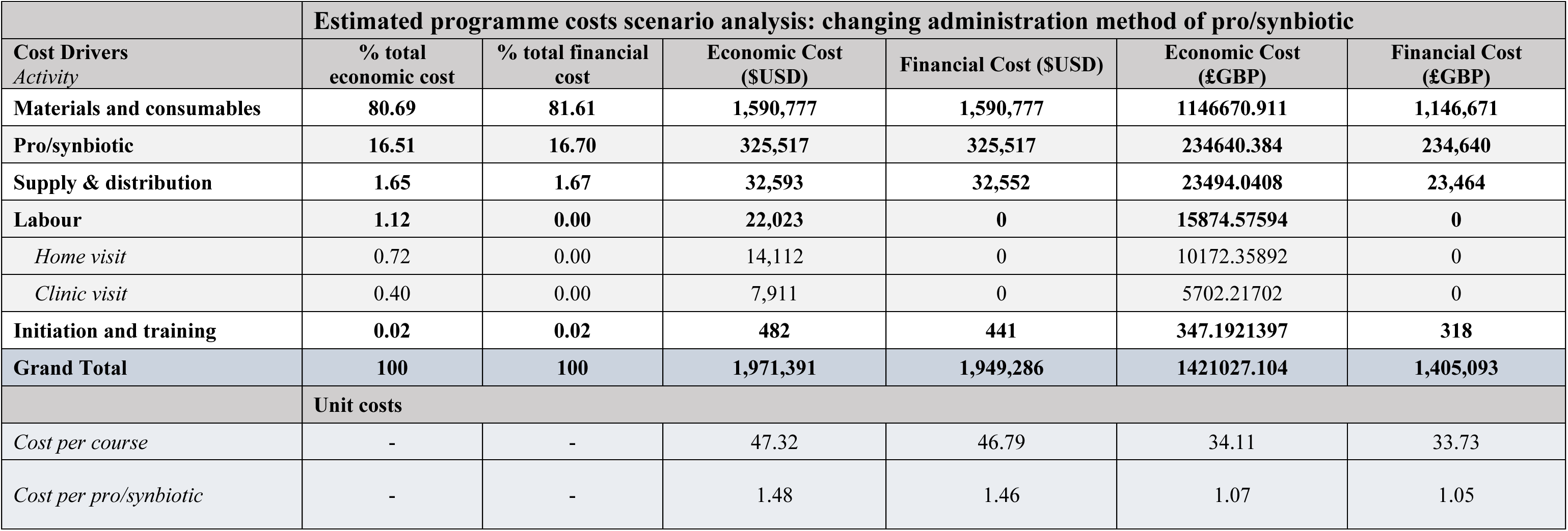
Estimated programme costs scenarios analysis: administration method.

**Supplementary Table 7:**
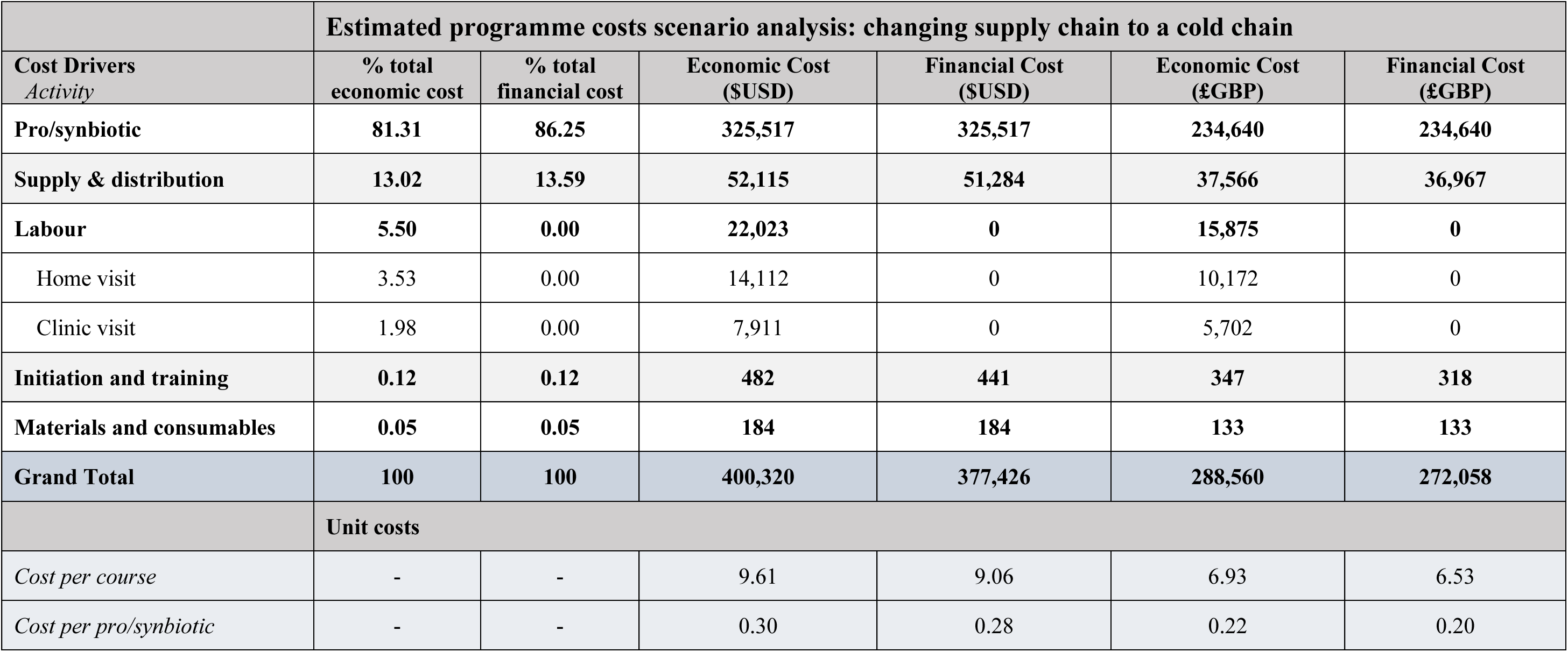
Estimated programme costs scenarios analysis: cold chain.

1. Republic of Kenya Ministry of Health (MoH), *Homa Bay County, Health at a Glance*. 2015.
2. Institute for Health Metrics and Evaluation (IHME). *Kenya - HomaBay*. 2019 13 August 2021]; Available from: http://www.healthdata.org/kenya-homabay.
3. County Innovation Challenge Fund (CICF), *Ubuntu-Afya Kiosks: An innovative partnership for extending quality healthcare services to underserved communities* 2019.
4. Republic of Kenya Ministry of Health (MoH), *Health Sector HRH Strategy 2014 – 2018*. 2014.

